# Frailty and hospitalization in older adults with type 2 diabetes in Vietnam: a focus on sex disparities

**DOI:** 10.1101/2025.08.04.25332930

**Authors:** Tan Van Nguyen, Dung Ngoc Truong, Wei Jin Wong, Mark Woodward, Tu Nguyen

## Abstract

**Background:** Older adults with diabetes often face multiple health challenges such as frailty. Frailty has been reported to be more common and severe in women than men, potentially due to a combination of biological, social and environmental factors. These sex-related differences in frailty may influence health outcomes, such as hospitalization rates.

**Aim:** This study sought to examine the prevalence of frailty in older adults with type 2 diabetes in an older population in Vietnam, and the association between frailty and hospitalizations, with a focus on sex disparities.

**Method:** An observational cohort study was conducted at two tertiary hospitals in Vietnam from November 2022 to June 2023. Patients aged 60 years or above with type 2 diabetes that visited the cardio-metabolic clinics during the study period were recruited. Frailty was measured using Fried’s frailty criteria. Logistic regression models were applied to examine the association between frailty and all-cause hospitalization over 6 months. Ratios of odds ratios (ORs) were computed to quantify the sex difference.

**Results:** There were 644 participants, with a mean age of 71.8 years (SD 7.6), and 30.0% were classified as frail. The prevalence of frailty in women was higher compared to men (31.3% vs 28.8%, p<0.001). The adjusted ORs of frailty on 6-month all-cause hospitalization were 2.71 (1.07 – 6.82) in women, and 1.92 (0.78 – 4.75) in men.

**Conclusion:** In this study, frailty was more prevalent in women and was associated with an increased risk of hospitalizations in women than in men. This study adds to the understanding of how frailty and sex influence health outcomes in older adults with diabetes, implying the need for sex-specific approaches in managing diabetes in older adults.

## Introduction

Older adults with diabetes often encounter a variety of age-related challenges that can complicate their management of this condition. These issues include frailty, multimorbidity, polypharmacy, and diabetes-related complications.^1-3^ The increasing recognition of frailty as a common geriatric condition among older adults also adds another layer of complexity to diabetes management.^3,4^ Frailty is a condition characterized by increased vulnerability to adverse health outcomes, often due to age-related declines in physiological reserves.^5^ Frailty and diabetes have been described as having a bidirectional relationship.^6^ Diabetes can accelerate the ageing processes and the development of frailty through mechanisms such as chronic inflammation, oxidative stress, and microvascular complications.^6^ On the other hand, frailty can hinder effective self-management of diabetes by affecting medication adherence, dietary plans, and physical activity, resulting in poor glycemic control and diabetic complications.^3,6,7^ Moreover, the variations in how frail individuals metabolize and respond to medications can greatly influence the efficacy and safety of glucose lowering therapies and cardiovascular protective medications.^7-11^ In older patients with diabetes, frailty can reduce quality of life, increase healthcare costs, and hospitalizations.^3,7^ Understanding the impact of frailty on hospitalizations among older patients with diabetes is crucial for optimizing care, reducing negative outcomes, and informing healthcare policies.^7^

Sex differences related to frailty, diabetes, and metabolism may also influence health outcomes in older adults with diabetes. Regarding frailty, there has been evidence that frailty occurs more frequently, and with greater severity, in women compared to men, which may be attributed to various biological, social, and environmental factors.^12,13^ Sex differences in diabetes have also been reported in many studies.^14,15^ Women with diabetes often face worse outcomes than men, due to socioeconomic factors, healthcare access, and biological differences.^14-16^ Women with diabetes were also reported to have a higher risk of cardiovascular complications compared to men.^17,18^ Biological sex influences both diabetes and frailty; however, the relationship between sex, frailty, and adverse outcomes in diabetes remains understudied. Understanding the sex differences in this context is critical for tailoring interventions, optimizing care, and reducing disparities in health outcomes between men and women.

In this study, we aimed to examine the prevalence of frailty in older adults with type 2 diabetes in an older population in Vietnam, and the association between frailty and hospitalizations, with a particular focus on sex disparities. Vietnam is an Asian country experiencing a concerningly rapid increase in diabetes, with approximately 5.8 million people with diabetes currently.^19,20^ By the year 2035, the prevalence of diabetes is expected to be 7.0%, while prediabetes is anticipated to be 15.7% in this country.^21^ However, there is limited evidence on the relationship between frailty and diabetes in older people in Vietnam.

## Methods

### Study design and population

An observational cohort study was conducted at the cardio-metabolic clinics of two urban hospitals (Thong Nhat Hospital and Gia Dinh Hospital) in Ho Chi Minh City from November 2022 to June 2023. Consecutive patients aged ≥ 60 years diagnosed with type 2 diabetes who visited these clinics during the study period were recruited. The exclusion criteria included: (1) having dementia or having a mental illness that can affect their ability to answer the study questionnaires, (2) having hearing impairments that hinder their ability to answer the study questionnaires, (3) having an acute illness or condition that require hospitalization in the next 24 hours, and (4) not being able to provide consent.

### Data collection

Data were collected from patient interviews and medical records. Information obtained included demographic characteristics, height, weight, medical history, duration of having diabetes (in years), and comorbidities. Educational status was defined using four levels of completion (primary school, high school, university/college, and postgraduate). Smoking was defined as current smoking (yes vs. no). Body mass index (BMI) was calculated from measured weight and height and classified into four groups: underweight (BMI < 18.5 kg/m^2^), ideal (BMI 18.5–22.9 kg/m^2^), overweight (BMI 23.0–24.9 kg/m^2^), and obese (BMI ≥ 25.0 kg/m^2^). Comorbidities were assessed using the Charlson Comorbidity Index (CCI), which estimates the 10-year risk of death associated with a combination of comorbidities. Based on the CCI, the severity of comorbidity was categorized as mild (CCI 1-2), moderate (CCI 3-4), and severe (CCI ≥ 5).^22^ Polypharmacy was defined as the use of five or more medications daily.^23^ Duration of diabetes was defined as the period from the initial diagnosis of type 2 diabetes to the time of the interview. HbA1c values were obtained from the latest measurement within the past three months. Poor glycaemic control was defined as HbA1c ≥7.0%.^24^

Frailty assessment: Frailty was defined by Fried’s frailty criteria,^5^ which includes 5 components: unintentional weight loss, weakness, exhaustion, slowness and low physical activity. Participants with three or more of these components were identified as being frail, those with one or two components were identified as prefrail, and those with none of these of components as robust. Weight loss was defined as an unintentional weight loss of ≥5% or 4.5 kg in the last year. Grip strength was measured with a hand dynamometer twice in the dominant hand and the highest value was used. Weakness was defined by a low grip strength, using the cut-offs suggested by Fried’s study^5^: in men, ≤29 kg if BMI ≤24.0 kg/m^2^, ≤30 kg if BMI 24.1 to 28.0 kg/m^2^, ≤32 kg if BMI >28.0 kg/m^2^; in women, ≤17 kg if BMI ≤23.0 kg/m^2^, ≤17.3 kg if BMI 23.1 to 26.0 kg/m^2^, ≤18 kg if BMI 26.1 to 29.0 kg/m^2^, and ≤21 kg if BMI >28.0 kg/m^2^. Exhaustion was defined by two questions from the Centre for Epidemiologic Studies Depression Scale (CES-D): In the last week “I felt that everything I did was an effort” and “I couldn’t get going”. Participants who answered “frequently” or “always” to at least one of these two questions were classified as having this criterion. Slowness was defined according to the time taken to complete the 4.6-metre walking test, with the following cut-offs: ≥ 6 seconds for men with a height ≤173 cm and women with a height ≤159 cm, ≥ 7 seconds for men taller than173 cm and women taller than 159 cm. The International Physical Activity Questionnaire (IPAQ) was used to measure vigorous and moderate activities, and walking in the previous 7 days. The metabolic equivalent task (MET, in minutes per week) of each category was calculated by multiplying the reported weekly minutes spent by the corresponding MET score (8 for vigorous activities, 4 for moderate activities and 3.3 for walking). The total score, measured in MET-minutes per week, was calculated by adding the values from these three activity categories (vigorous activity, moderate activity, and walking). This score was then converted to kilocalories (Kcal). Low physical activity was defined as <383 Kcal/week for men and < 270 Kcal/week for women.

### Outcomes

The study outcome was all-cause hospitalization over 6 months. Hospitalization information was obtained from patient medical records and by making phone calls to the participants or their caregivers.

The studies were approved by the Ethics Committee of the University of Medicine and Pharmacy at Ho Chi Minh City (Reference Number 934/DHYD-HDDD, dated 24/11/2022). Informed consent was obtained from all participants.

### Statistical analysis

Participant characteristics are presented as mean and standard deviation (SD) for continuous variables, and frequencies and percentages for categorical variables. Comparisons among groups were conducted using chi-square tests or Fisher’s exact tests for binary variables, and Student’s t-tests or Mann-Whitney U tests for continuous variables.

Logistic regression models were applied to examine the association between frailty and all-cause hospitalization over 6 months. These models were adjusted for age, marital status, education, Charlson Comorbidity Index, HbA1c level and years of having diabetes. Frailty was treated as a binary variable (frail/non-frail) in these models. We also conducted sensitivity analyses with frailty as a continuous score and as an ordinal variable with three levels (robust, prefrail, frail). Results are presented as odds ratios (ORs) and 95% confidence intervals (CIs), and an interaction term was added to the models to obtain the women-to-men ratios of ORs and 95% CIs, which were used to quantify how the ORs differed between the sexes.^25^ P values <0.05 were considered statistically significant. Data were analysed in SPSS Statistics 29.0.

## Results

A total of 644 participants (340 men, 304 women) with type 2 were recruited. They had a mean age of 71.8 (SD 7.6) years. The mean duration of having diabetes was 10.8 (SD 7.4) years, higher in men (11.8 years, SD 7.8) compared to women (9.7 years, SD 6.6) (Table 1). Women were more likely to be widowed (26.6% vs. 17.1% in men, p=0.032) and had lower education than men. Men had higher prevalence of smoking (51.8% vs. 1.0% in women, p<0.001), polypharmacy (80.3% vs. 73.4% in women, p=0.037), ischemic stroke (2.4% vs. 0.3% in women, p=0.040), and heart failure (2.4% vs. 0% in women, p=0.008).

**Table 1.** Participants characteristics.

| Characteristic | All participants<br>(N = 644) | Men<br>(n=340) | Women<br>(n=304) | p value<br>comparing<br>sexes |
| --- | --- | --- | --- | --- |
| Age, in years (mean, SD) | 71.8 (7.6) | 71.4 (7.9) | 72.3 (7.2) | 0.110 |
| Working status |  |  |  |  |
| Retired | 617 (95.8) | 330 (97.1) | 287 (94.4) | 0.094 |
| Working | 27 (4.2) | 10 (2.9) | 17 (5.6) |  |
| Marital status |  |  |  |  |
| Married | 479 (74.4) | 268 (78.8) | 211 (69.4) | 0.032 |
| Never married | 18 (2.8) | 10 (2.9) | 8 (2.6) |  |
| Divorced/separated | 8 (1.2) | 4 (1.2) | 4 (1.3) |  |
| Widowed | 139 (21.6) | 58 (17.1) | 81 (26.6) |  |
| Education |  |  |  |  |
| Primary school or less | 56 (8.7) | 12 (3.5) | 44 (14.5) | <0.001 |
| High school | 303 (47.0) | 136 (40.0) | 167 (54.9) |  |
| College/University | 227 (35.2) | 151 (44.4) | 76 (25.0) |  |
| Higher education | 58 (9.0) | 41 (12.1) | 17 (5.6) |  |
| Body mass index, kg/m<sup>2</sup> |  |  |  |  |
| Underweight (<18.5) | 17 (2.7) | 5 (1.5) | 12 (3.9) | 0.130 |
| Normal (18.5 – 22.9) | 343 (53.5) | 174 (51.6) | 169 (55.6) |  |
| Overweight (23.0 – 24.9) | 170 (26.5) | 95 (28.2) | 75 (24.7) |  |
| Obese (≥25.0) | 111 (17.3) | 63 (18.7) | 48 (15.8) |  |
| Smoking (yes vs. no) | 179 (27.8) | 176 (51.8) | 3 (1.0) | <0.001 |
| Duration of having diabetes, in years (median and range) | 10.0 (1 – 40) | 10.0 (1 – 40) | 9.5 (1 – 36) | 0.003 |
| Total number of medications (mean, SD) | 5.6 (1.4) | 5.7 (1.4) | 5.4 (1.4) | 0.012 |
| Polypharmacy | 496 (77.0) | 273 (80.3) | 223 (73.4) | 0.037 |
| Charlson Comorbidity Index (mean, SD) | 2.5 (1.1) | 2.5 (1.2) | 2.4 (1.0) | 0.255 |
| Details of comorbidities |  |  |  |  |
| Dyslipidemia | 630 (97.8) | 332 (97.6) | 298 (98.0) | 0.742 |
| Hypertension | 615 (95.5) | 327 (96.2) | 288 (94.7) | 0.379 |
| Chronic kidney disease | 104 (16.1) | 64 (18.8) | 40 (13.2) | 0.051 |
| Peripheral artery | 71 (11.0) | 34 (10.0) | 37 (12.2) | 0.380 |

| disease |  |  |  |  |
| --- | --- | --- | --- | --- |
| Coronary heart disease | 11 (1.7) | 7 (2.1) | 4 (1.3) | 0.552 |
| Ischemic stroke | 9 (1.4) | 8 (2.4) | 1 (0.3) | 0.040 |
| Heart failure | 8 (1.2) | 8 (2.4) | 0 (0) | 0.008 |

Continuous data are presented as mean (SD-standard deviation) or median (range).
Categorical data are shown as n (%).

### Sex differences in frailty and its components

Overall, 30.0% of the participants were classified as frail, 45.5% as prefrail, and 24.5% as non-frail. Women had significantly higher prevalence of prefrailty (52.0% vs. 39.7%) and frailty (31.3% vs. 28.8%) compared to men (p<0.001). Among the five components of Fried’s frailty criteria, women had a significantly higher prevalence of having low grip strength (74.3% vs. 55.3%) and low physical activity (56.3% vs. 48.5%) than men. (Table 2)

**Table 2.** Frailty and the components of Fried’s frailty criteria among women and men.

|  | All participants<br>(n=644) | Men<br>(n=340) | Women<br>(n=304) | p value<br>comparing<br>sexes |
| --- | --- | --- | --- | --- |
| Fried's frailty score | 1.9 (1.4) | 1.7 (1.5) | 2.1 (1.3) | 0.002 |
| Frailty |  |  |  |  |
| Non-frail | 158 (24.5) | 107 (31.5) | 51 (16.8) | <0.001 |
| Pre-frail | 293 (45.5) | 135 (39.7) | 158 (52.0) |  |
| Frail | 193 (30.0) | 98 (28.8) | 95 (31.3) |  |
| Fried's frailty components |  |  |  |  |
| Weight loss | 48 (7.5) | 28 (8.2) | 20 (6.6) | 0.424 |
| Low grip strength | 414 (64.3) | 188 (55.3) | 226 (74.3) | <0.001 |
| Slowness | 259 (40.2) | 126 (37.1) | 133 (43.8) | 0.084 |
| Exhaustion | 171 (26.6) | 86 (25.3) | 85 (28.0) | 0.444 |
| Low physical activity | 336 (52.2) | 165 (48.5) | 171 (56.3) | 0.050 |

Data are shown as n (%).

### Sex differences in the impact of frailty on 6-month hospitalization

During the 6-month follow-up, 13.8% of the participants were admitted to hospitals. Among women, the hospitalization rates were 25.3% in the frail vs 9.6% in the non-frail, p <0.001. Among men, the hospitalization rates were 21.4% in the frail vs 9.9% in the non-frail, p = 0.005. (Figure 1)

**Figure 1.**
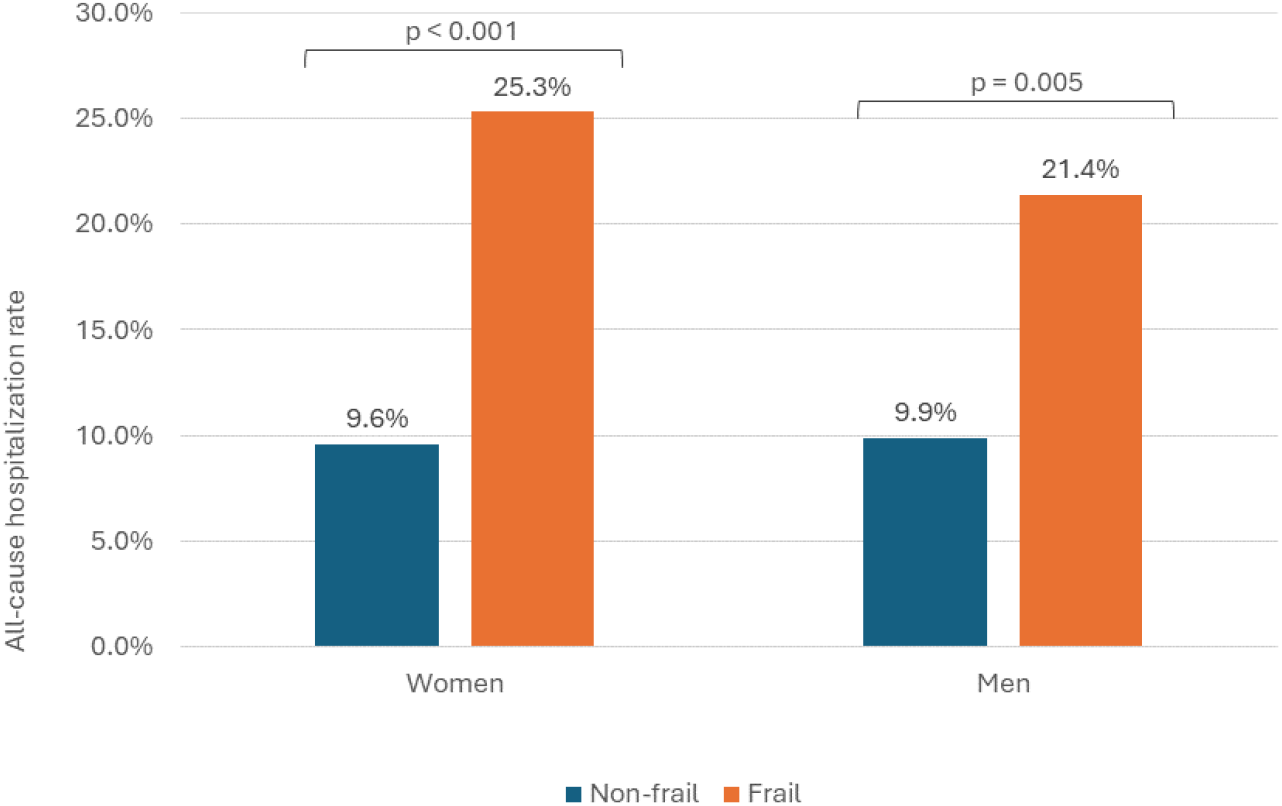
Hospitalization rate at 6 months by sex and frailty.

In the multivariable logistic regression models adjusted for age, marital status, education, Charlson Comorbidity Index, HbA1c level, years of having diabetes, the adjusted ORs of frailty on 6-month all-cause hospitalization were 2.71 (1.07 – 6.82) in women, 1.92 (0.78 – 4.75) in men, women to men ratio of ORs 1.41 (0.39 – 5.15). (Table 3)

**Table 3.** Sex differences in the association of frailty with 6-month all-cause hospitalization among older participants with diabetes.

|  | Odds Ratio (95%CI) |  | Women-to-Men<br>Ratio of Odds<br>Ratios (95%CI) |
| --- | --- | --- | --- |
|  | Women | Men |  |
| Unadjusted models |  |  |  |
| Binary frailty (yes vs. no) | 3.19 (1.66 – 6.14) | 2.48 (1.31 – 4.70) | 1.29 (0.52 – 3.21) |
| Three levels of frailty |  |  |  |
| Robust | 1 | 1 |  |
| Prefrail | 3.15 (0.71 – 14.07) | 2.06 (0.82 – 5.16) | 1.53 (0.26 – 8.83) |
| Frail | 8.28 (1.87 – 36.60) | 3.90 (1.58 – 9.64) | 2.12 (0.37 – 12.10) |
| Fried's frailty score (1 to 5) | 1.63 (1.26 – 2.10) | 1.37 (1.11 – 1.70) | 1.19 (0.85 – 1.66) |
| Adjusted* models |  |  |  |
| Binary frailty (yes vs. no) | 2.71 (1.07 – 6.82) | 1.92 (0.78 – 4.75) | 1.41 (0.39 – 5.15) |
| Three levels of frailty |  |  |  |
| Robust | 1 | 1 |  |
| Prefrail | 4.98 (0.61 – 40.74) | 2.38 (0.80 – 7.06) | 2.09 (0.20 – 22.30) |
| Frail | 12.40 (1.35 – 113.94) | 3.85 (1.09 – 13.67) | 3.22 (0.25 – 41.37) |
| Fried's frailty score (1 to 5) | 1.65 (1.14 – 2.39) | 1.32 (0.97 – 1.81) | 1.25 (0.74 – 2.03) |

\*Adjusted for age, marital status, education, Charlson Comorbidity Index, HbA1c level and duration of diabetes

Sensitivity analyses with frailty as a continuous score and as an ordinal variable yielded similar results. For every one-point increase in Fried’s frailty score, the adjusted ORs of frailty on 6-month all-cause hospitalization were 1.65 (1.14 – 2.39) in women, 1.32 (0.97 – 1.81) in men, women to men ratio of ORs 1.25 (0.74 – 2.03). When frailty was categorized into three levels, compared to robust individuals, the odds of hospitalization significantly increased in the frail: adjusted ORs 12.40 (1.35 – 113.94) in women, 3.85 (1.09 – 13.67) in men, women to men ratio of ORs 3.22 (0.25 – 41.37), but no significant differences were observed between the prefrail and the robust, in both women and men. (Table 3)

## Discussion

In this study of 644 older participants with type 2 diabetes, the prevalence of frailty and prefrailty was higher in women, and the impact of frailty on hospitalization was also more significant in women than in men.

Our finding aligned with findings from studies in older patients with diabetes. In another multicentre study of 638 older adults with type 2 diabetes in Vietnam, pre-frail and frailty was also reported to be higher in women vs men (pre-frail: 56.7% vs 51.7%, frailty: 29.9% vs 26.8%).^26^ In a study of 2403 of community-dwelling older adults from the Korean Frailty and Aging Cohort Study, among the subgroup of participants with diabetes, the prevalence of pre-frailty (60.9% vs 54.2%) and frailty (9.5% vs 5.5%) was higher in women compared to men.^27^

We found that women were more likely to exhibit low grip strength and low levels of physical activity. Grip strength is a non-invasive method used to assess muscle weakness in clinical practice.^28^ Out results also align with findings from the Swedish Adoption/Twin Study of Aging, which highlighted sex differences in grip strength.^29^ In that study, researchers followed grip strength performance from the age of 50 up to the last years of life of 849 participants who were 50–88 years of age at baseline, with a follow-up period of 22 years. They identified an age difference of 5 years, with women experiencing a decline in grip strength starting at age 67 and men starting at age 72. The average grip strength level for women at age 67 was 21.6 kg, with a decline of 0.19 kg/year between ages 50 and 67 and 0.45 kg/year from ages 67 to 96. In comparison, men had an average grip strength of 36.3 kg at age 72, declining by 0.51 kg/year between 50 and 72 years of age, and 0.95 kg/year from ages 72 to 96. That study also revealed sex differences in the pattern of risk factors associated with grip strength decline: for women, the risk factors appeared to be more lifestyle related (eg. smoking, stress, depression), while for men the risk factors were more associated with physical conditions (e.g. physical activity, chronic health disorders, and mean arterial pressure).^29^ These findings emphasize the potential for targeted frailty intervention aimed at women. When designing interventions to reduce frailty, it is important to consider sex-specific issues. For example, prioritizing customized strength training and encouraging women to enhance their physical activity can be effective strategies. The multifaceted roles women often assume as caregivers can inadvertently restrict their capacity to engage in activities that promote physical activities and self-care.

There are several potential mechanisms for the observed sex differences on the impact of frailty on hospitalization in our study. Sex differences in pharmacokinetics and pharmacodynamics can affect the effectiveness and adverse side effects of glucose lowering therapies.^30^ Women may experience more side effects from certain antidiabetic medications, influencing treatment adherence, all of which can lead to an increased risk of hospitalizations.^31^ Disparities in healthcare access may also contribute to a difference in the impact of frailty on hospitalization. Women, particularly in low-resource settings, may face barriers to healthcare access, delaying diagnosis and treatment of diabetes-related conditions.^32^ Women are more likely to prioritize family or caregiving responsibilities over self-management, leading to poorer adherence to diabetes care plans compared to men.^33^ These sex differences may also be partly due to the loss of estrogen’s cardioprotective effects in older women with diabetes.^34-36^

Our findings suggest that routine assessment of frailty should be performed for older adults with type 2 diabetes. Clinicians should consider sex differences when assessing frailty, and women may require more proactive screening for frailty. Additionally, our study underscores the need for further research into why frailty poses a higher hospitalization risk for women. This research could help inform sex-specific healthcare policies or guidelines. Women with frailty may need targeted preventive measures, such as such as exercise programs (e.g., resistance training), nutritional support, polypharmacy management, home-based care, telehealth monitoring, or community support programs, to reduce hospital admissions.

### Strengths and limitations

To the best of our knowledge, this is the first study examining sex differences in how frailty affects the risk of hospitalization in older adults with type 2 diabetes. The study comprised high-quality, multi-centre data and clinical relevance. However, the study has several potential limitations. We did not collect data on income, nutrition intake, medication use (including glucose lowering therapies) and medication adherence, factors that may serve as potential confounders for the relationship between frailty and hospitalizations. Future studies would benefit from including these parameters to provide a more comprehensive analysis and to enhance the robustness of the findings. Previous studies in adults with cardiovascular diseases in Vietnam reported that women had lower income compared to men.^37,38^ This disparity may lead to reduced access to healthcare and glucose lowering medications for women. The study participants were recruited from two major hospitals in Ho Chi Minh City, an urban city in Vietnam. Therefore, the findings should be interpreted within this context and may not be generalisable to other regions, such as rural areas. Finally, the study was underpowered to be able to detect important sex differences in associations between frailty and hospitalisation. Further analyses, and meta-analyses, are required to produce robust, generalisable, results.

## Conclusions

In this study in older adults with type 2 diabetes, frailty was more common in women and was associated with an increased risk of hospitalizations in women than in men. This study contributed to the understanding of how frailty and sex influence health outcomes in older adults with diabetes, and informs the development of targeted interventions to improve health outcomes for this population. These findings highlight the need for sex-specific approaches in managing diabetes in for older adults. This knowledge could also aid in developing future care strategies that are specific to each gender.

## Data Availability

All data produced in the present study are available upon reasonable request to the authors

## Acknowledgements

We thank all the participants for their participation in this study.

## Author Contributions

TVN and TN conceptualized the analysis. TVN and TN conducted the statistical analyses and led the manuscript writing. All authors were involved in data interpretation. The manuscript was revised for important scientific content by all authors. All authors approved the final version of the manuscript.

